# A protocol for high-dose quadrivalent influenza vaccine effectiveness in the community and long-term care facilities using electronic health records

**DOI:** 10.1101/2024.11.08.24316963

**Authors:** Patrícia Soares, Verónica Gomez, Vânia Gaio, João Almeida Santos, Ana Paula Rodrigues, Ausenda Machado

**Author notes:** **Funding:** This work was partially supported by FCT – Fundação para a Ciência e Tecnologia, I.P. by project reference CEECINST/00049/2021/CP2817/CT0001 and DOI identifier 10.54499/CEECINST/00049/2021/CP2817/CT0001.

## Abstract

Since the 2022-2023 season in Portugal, a high-dose quadrivalent influenza vaccine is freely available for individuals living in long-term care facilities (LTCF). In 2024-2025, vaccination was extended to community-dwelling individuals aged ≥85 years. Given the scarcity of reported high-dose influenza vaccine effectiveness (IVE) estimates for this population, this study aims to estimate the high-dose relative and absolute IVE.

A retrospective cohort study using data from electronic health records databases (EHR) will be implemented, using two cohorts, one of individuals vaccinated with influenza vaccine (to estimate relative IVE) and another of individuals eligible for the high-dose quadrivalent influenza vaccine (to estimate absolute IVE). We will consider two subgroups for both cohorts: individuals living in LTCF and community-dwelling individuals aged ≥85. We will use a fixed cohort approach, defining the eligible population by age at the vaccination campaign(s) start and living status. The outcomes are based on the primary cause of hospital admission. The reference population database will be defined by linking EHR on vaccination, comorbidities, and hospitalisations using a unique identifier through a deterministic data linkage procedure, and influenza vaccination status will be assessed retrospectively. We will use Cox proportional hazards regression models to estimate the hazard ratio (HR), considering as event the first hospitalisation due to influenza-like-illness and as exposure the vaccination status. IVE will be estimated as one minus the confounder-adjusted HR of vaccinated with the high-dose quadrivalent influenza vaccine vs vaccinated with standard dose (to estimate relative IVE) or unvaccinated (to estimate absolute IVE).

While challenges such as EHR constraints and potential reporting bias pose limitations, using routinely collected data has successfully estimated COVID-19 VE and enables precise monitoring of VE with higher representativeness. The results of this study will inform the Health Ministry on the future influenza vaccine programme in Portugal.

## Background

In Portugal, as in most European countries, influenza seasonal immunisation is recommended annually to high risk individuals, which include, among others, elderly individuals (age stratification varies across seasons, i.e., aged ≥60 or aged ≥65 years) and those who reside in long-term care facilities (LTCF)(1).

Adults aged 65 or more are more prone to lower respiratory tract infections, including pneumonia, bronchitis, and tracheobronchitis, due to specific factors, such as genetic polymorphisms, chronic immunocompromised conditions, age and correspondent age-related comorbidities, that may compromise the individual capacity to produce an adequate immune response(2,3). These infections are associated with considerable morbidity and can lead to hospitalisation, especially in frail older adults, such as those residing in LTCF (4). There is a potentially different risk of exposure in older community-dwelling individuals due to the greater number of contacts. Additionally, LTCFs significantly contribute to the variability in exposure to the disease between individuals, resulting in greater individual susceptibility to severe outcomes (4,5).

Vaccination is widely considered the most effective intervention against influenza and its associated complications (5). However, influenza vaccination strategies might target individuals who, although at high risk of influenza complications, may have impaired capacity for developing effective vaccine-induced immunity. Given the specificities of individuals in LTCF, the effect of influenza vaccination in this population needs further investigation (2,5). A high-dose quadrivalent influenza vaccine was developed to provide greater protection and better prevent influenza-related complications than the standard dose influenza vaccine.

This high-dose quadrivalent vaccine presented higher efficacy in a randomised clinical trial versus a standard-dose influenza vaccine for preventing laboratory-confirmed influenza illness in adults aged 65 or more (3). Systematic reviews and meta-analyses, which included randomised and observational studies, assessed the effectiveness of high-dose inactivated influenza vaccines versus standard-dose influenza vaccines against influenza-related outcomes in 65 or more years old individuals (6,7). The results support the evidence on the effectiveness of high-dose inactivated influenza vaccines compared to standard-dose influenza vaccines in preventing severe influenza outcomes in 65 or more years old individuals, with additional support from observational data (7). Regarding severity-related outcomes, the high-dose quadrivalent influenza vaccine was more effective than the standard-dose influenza vaccine in preventing influenza-related hospitalisations, with a relative influenza vaccine effectiveness (rIVE) of 11.2% (7.4%-14.8%) for all seasons. Despite the important evidence of the effectiveness of the influenza vaccine (IVE) of the high-dose inactivated influenza vaccines for individuals aged 65 or older, only three studies in nursing homes were included (6). Review studies indicated a need for further real-world population-based studies of high-dose influenza vaccines. Hence, it remains necessary to assess the clinical effectiveness of this vaccine in older adults living in LTCF (7–9).

In Portugal, since the 2022-2023 season (1,10), the high-dose quadrivalent influenza vaccine is available exclusively for individuals aged 65 or more living in LTCF, free of charge. In the 2023-24 season, the high-dose influenza vaccine became available to community-dwelling individuals aged 65 or more in community pharmacies, with co-payment if the patient presents a prescription (11).

Given the scarcity of reported high-dose IVE estimates for this population and the Portuguese strategy of vaccinating institutionalised individuals aged 65 or more, it is crucial to evaluate the vaccine’s effect on preventing severe outcomes in this age group of the Portuguese population. Thus, the objective of this study is to estimate the rIVE of the high-dose quadrivalent influenza vaccine against hospital admission due to influenza-like-illness in individuals vaccinated with any influenza vaccine and the absolute IVE of the high-dose quadrivalent influenza vaccine against hospital admission due to influenza-like-illness in individuals eligible for the high-dose quadrivalent vaccine, using data routinely collected in electronic health records (EHR) in mainland Portugal.

## Methods

### Study design and population

A retrospective cohort study using data collected routinely in EHR databases will be implemented. For the estimation of rIVE, only vaccinated individuals will be included, either with the high-dose or the standard influenza vaccine. For the estimation of absolute IVE, individuals eligible for the high-dose influenza vaccine during the study period will be included. For both cohorts, we will consider two subgroups: community-dwelling individuals aged ≥85 years and individuals living in LTCF. The cohorts are represented in Figure 1, with panel A representing rIVE and panel B absolute IVE.

**Figure 1:**
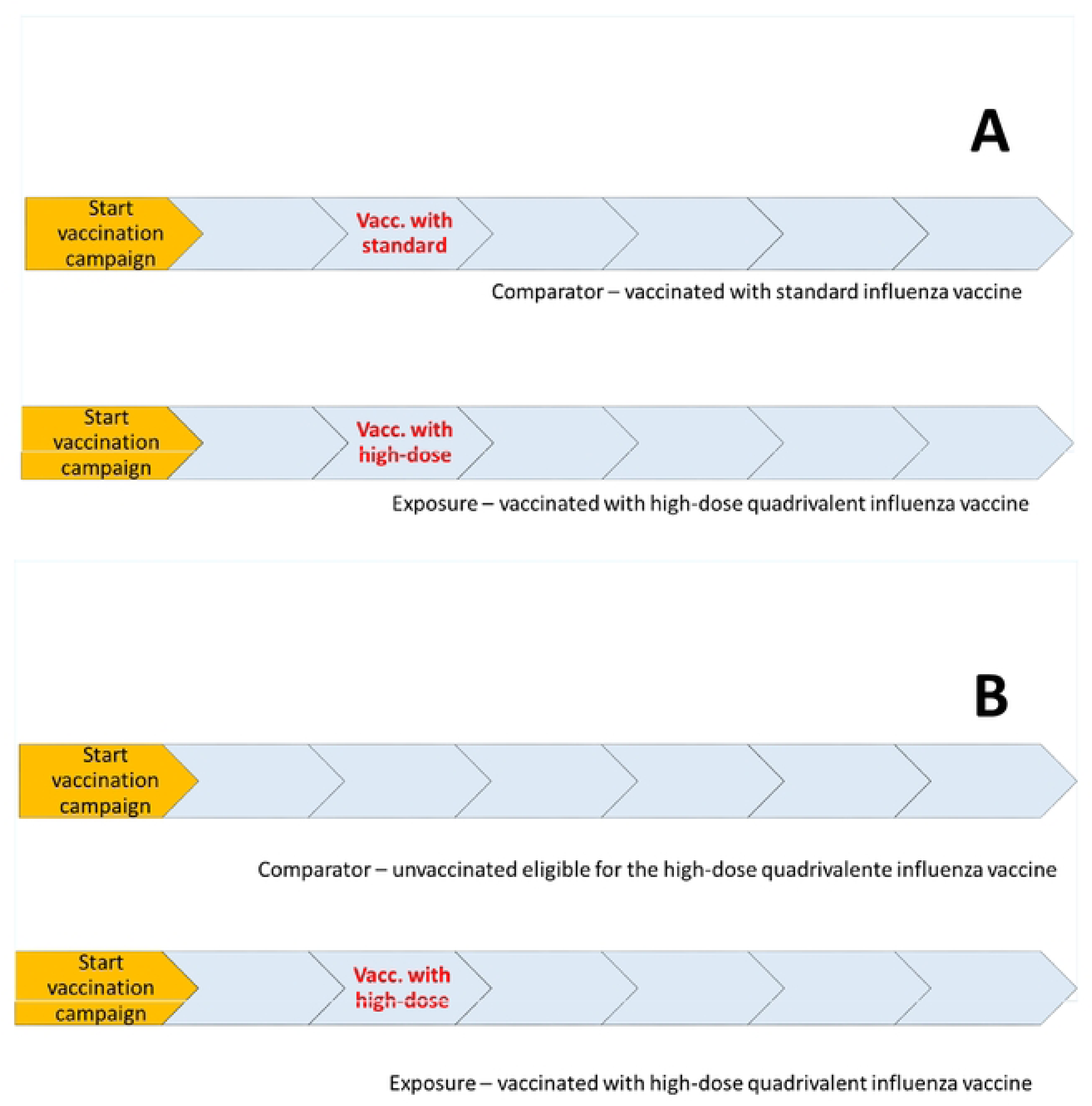
Diagram of the two cohorts designed in this study. A – cohort of vaccinated individuals to measure relative influenza vaccine effectiveness, B – cohort of individuals eligible for the high-dose influenza vaccine to measure absolute influenza vaccine effectiveness. For the two cohorts, we will consider two subgroups: community-dwelling individuals aged ≥85 years and individuals living in LTCF.

We will use a fixed cohort approach, defining the eligible population by their age at the start of the vaccination campaign(s) and living status. Table 1 describes the connection between the cohorts and the objectives.

**Table 1.**
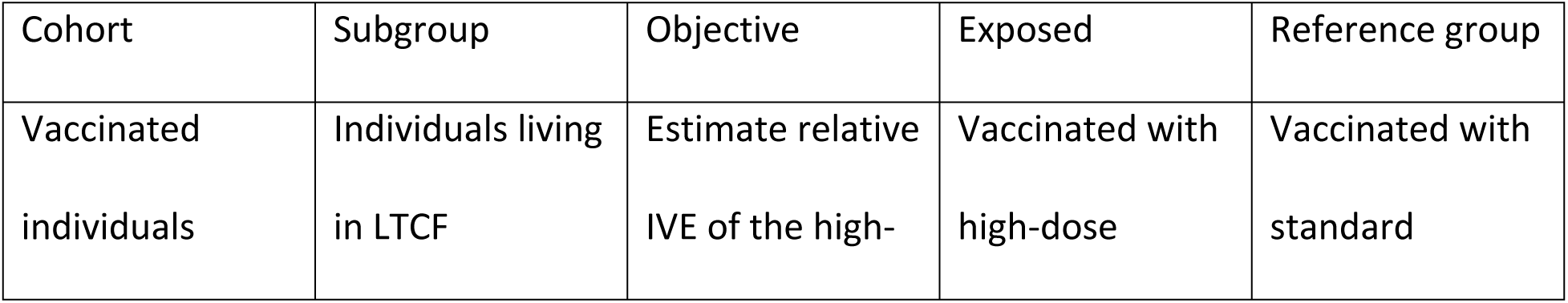
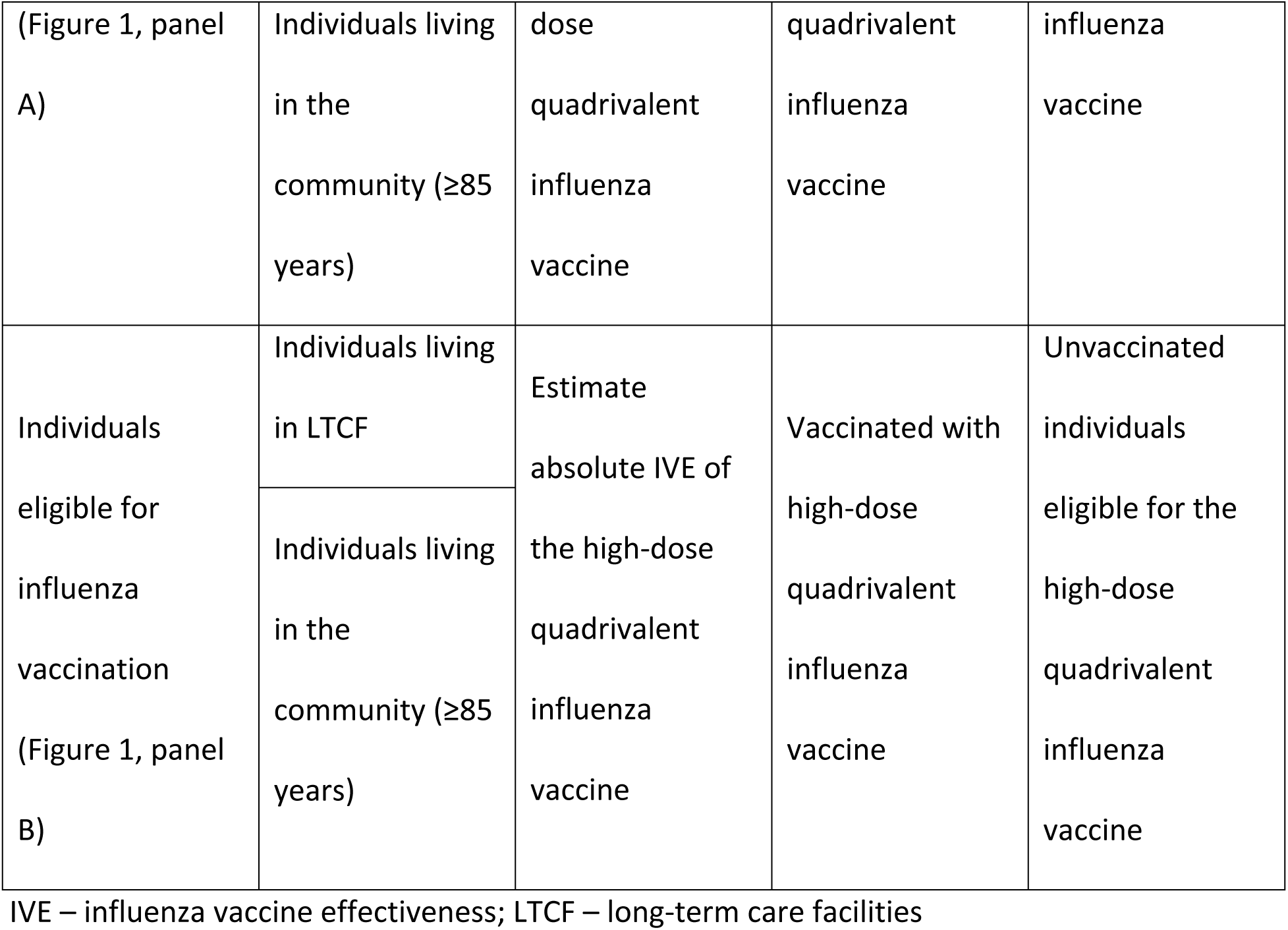
Summary of the study design with the different cohorts, objectives and exposed and unexposed groups.

### Inclusion and exclusion criteria

The study population includes all registered individuals without contraindications for influenza vaccination residents in mainland Portugal. Only vaccinated individuals with the high-dose quadrivalent or standard influenza vaccine living in LTCF or community-dwelling individuals aged 85 or older will be included to estimate rIVE. Whereas, to estimate absolute IVE, only eligible individuals for the high-dose influenza vaccine living in LTCF or community-dwelling individuals aged 85 or older will be included.

We will exclude individuals with inconsistent or missing data on vaccination (e.g. any vaccination date unknown and any vaccine brand unknown), individuals who received any vaccine brand not approved by the European Medicines Agency (EMA), and individuals vaccinated before the study started.

### Study setting and period

To estimate rIVE, the study population will include individuals vaccinated with the high-dose or standard influenza vaccine in mainland Portugal between the 2022/2023 and 2024/2025 seasons. When applicable, we will analyse each season (2022/2023, 2023/2024, 2024/2025) individually, starting after the implementation of each influenza vaccination campaign and ending nine months thereafter.

To estimate absolute IVE, the study population will include individuals for whom the high-dose influenza vaccine has been recommended in mainland Portugal for the 2024/2025 season. The study period will start after the implementation of the influenza vaccination campaign period and end nine months thereafter.

### Vaccination status

To estimate rIVE, the vaccination status will be assessed retrospectively, and the time since vaccination will be estimated. The exposure of interest will be to receive the high-dose quadrivalent influenza vaccine, and the comparison group will be composed of those who received the standard-dose influenza vaccine.

To estimate absolute IVE, the vaccination status will be assessed as a time-changing variable according to the following classification:

- Unvaccinated: person-time of individuals without any quadrivalent influenza vaccine in the season.
- Vaccinated: person-time of individuals who received a high-dose quadrivalent influenza vaccine dose during the study period. The completion status is achieved 14 days after administration of the dose.

### Time since vaccination

If the sample size allows it, the time since completion of the high-dose influenza vaccine will be analysed as a secondary objective. For the rIVE estimation, time since vaccination will be considered a confounder to add to the model. For the absolute IVE estimation, time since vaccination will be calculated at each time point and classified into the following categories:

- From time 14 days to ≤89 days after time 0 (i.e., <13 weeks, approximately 3 months);
- 90 to 179 days after time 0 (i.e., ≥13 weeks and <26 weeks, approximately 3 to 6 months);
- 180 to 272 days (i.e., ≥26 weeks and <39 weeks, approximately 6 to 9 months).

### Outcome

The outcomes of interest are defined based on the primary cause of admission to a hospital, coded according to ICD-10: J11 (Influenza with pneumonia), J12 (Viral pneumonia) and J18 (Pneumonia of unknown aetiology).

As a secondary outcome, we will also analyse cardiorespiratory hospitalisations as a primary cause of admission, with a secondary diagnosis of pneumonia (flu, viral or unknown aetiology). Each of these outcomes will be analysed separately. The outcome date will correspond to the hospitalisation date. Details on the ICD-10 codes are available in Supplementary Table S1.

### Stratification variables

If possible, and when applicable, rIVE will be estimated for each seasonal campaign.

### Potential confounding variables for adjustment

A set of variables will be used to account for confounding bias. Sociodemographic variables, such as sex, age, health administrative region, and the European Deprivation Index, will be used as potential confounding factors. We will also consider the vaccination against COVID-19 in the previous season to account for health-seeking behaviours. The presence of chronic conditions, such as diabetes, asthma, or other chronic respiratory diseases, cancer, chronic renal disease, hypertension or other chronic cardiovascular disease, obesity, chronic hepatic disease, neuromuscular disease, and immunodeficiency, will also be included as potential confounding factors.

### Data sources

The study will use routinely collected data from various population health registries. Each database will contain a unique identifier for each individual to allow data linkage between databases.

The National Health Service User (NHSU) dataset is the reference population database and includes individual records of the target study population. For this study, we will only consider individuals who had contact with the NHSU healthcare system three years before the start of the study period.

Vaccination registry or vaccination record databases with individual data, including influenza and vaccine brand dates, will be provided by the National Vaccination Registry – VACINAS.

Regarding the outcome, the National Hospital Discharges Registry will be used to assess hospitalisation information. We will also use data on mortality, which will be extracted from the National Death Registry, with the respective date of death.

### Construction of the cohort

#### Identification of individuals and characteristics at baseline

The reference population database will be linked with the electronic databases on vaccination, comorbidities and/or health-seeking behaviours, hospitalisations and other vital registries, using the unique identifiers through a deterministic data linkage procedure (no random component in the linkage procedure). Individuals will enter the study in their corresponding vaccination status group based on the data available in the vaccination registry.

Variables to be measured at baseline include age, sex, health administrative region, comorbidities, and other socioeconomic or health-seeking behaviour variables that will be used to adjust IVE estimates to stratify or account for confounding.

#### Time-changing characteristics (to estimate absolute IVE)

Vaccination status and time since vaccination will be assessed, and individuals will be classified into the same or updated vaccination status daily, generating a new record in the dataset for each new assessment. Person-time exposure between 0 and 13 days after vaccination will be excluded from the analysis.

#### Identification of outcomes during follow-up

Information for identifying outcomes and the dates when they occurred will be obtained by data linkage between the cohort built previously and the databases containing information on the respective outcomes. Outcome classification for each individual will be assessed from the start of the vaccination campaign.

#### Censoring events

All individuals will be followed from the start of the vaccination campaign until:

- Hospitalisation date (corresponding to the event date);
- At the end of the study (nine months after the start of the vaccination campaign);
- On the date of death (any cause).

### Analysis plan

#### Description of the sample selection

The total number of individuals fulfilling the inclusion criteria at the study baseline will be calculated for each cohort. The number and proportion of individuals excluded after applying each selection criteria will be recorded.

#### Description of the study population

The number of persons, total person-time of follow-up, and the number of events by vaccination status will be calculated. Distribution of the number of persons and total person-time of follow-up will be described by baseline variables in each vaccination status group considered in the study. To estimate the total number of persons, we will consider the vaccination status group at the end of each person-time follow-up (to estimate absolute IVE).

The proportion of the missing data will be used to determine if each specific variable can be included in the model and how (e.g., missing could eventually be included in the model as a category). Imputation to address missingness is not planned as a means for increasing data quality.

### Estimation of the vaccine effectiveness

A complete case analysis will be performed considering all variables in the final model to estimate adjusted-confounding IVE.

We will use Cox proportional hazards regression models to estimate the hazard ratio (HR), considering the event the first hospitalisation due to influenza-like-illness and as exposure the vaccination status. The crude HR of vaccinated with the high-dose influenza vaccine vs vaccinated with the standard dose (rIVE) or unvaccinated (absolute IVE) will be estimated for each outcome of interest during the study period, without adjusting for other factors or covariates.

We will consider two sets of confounding factors. First, we will estimate partially adjusted HR, adjusting by age, sex and country region. Age will be modelled using a restricted cubic spline, with knots specified according to Harrell (12). Second, a fully adjusted HR estimate will be produced by adjusting variables related to socioeconomic condition, comorbidities and health-seeking behaviour. IVE will be estimated as one minus the confounder-adjusted HR of vaccinated vs vaccinated with the standard dose (rIVE) or unvaccinated (absolute IVE) for each outcome of interest.

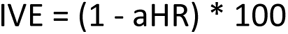

To estimate relative IVE, the time since vaccination will be modelled using a restricted cubic spline, with knots specified at zero and 15 days and then at the 40^th^ and 90^th^ percentile. To estimate absolute IVE for the time since vaccination (secondary objective), IVE will be estimated by comparing the hazard rate of the outcome in individuals vaccinated with the high-dose influenza vaccine for each class of the time since vaccination - 14 to 89 days, 90 to 179 days and 180 to 272 days (exposed group), in comparison with the outcome hazard rate in unvaccinated individuals (reference group).

### Propensity score matching

Individual characteristics might influence vaccination status, leading to systematic differences between those vaccinated with different vaccines and those vaccinated (rIVE) and unvaccinated (absolute IVE). Propensity score matching is a statistical technique used to reduce selection bias in observational studies by creating a matched set of treated (vaccinated with high-dose influenza vaccine) and untreated (vaccinated – rIVE and unvaccinated – absolute IVE) individuals with similar propensity scores. The first option will be matching without replacement, 1-to-1. However, considering the coverage vaccination, this ratio might need to be reviewed.

Several assumptions should be met to ensure that conditioning on the propensity score will lead to unbiased estimates of average treatment effects. The first one is that there should not be unmeasured confounders, i.e. the variables affecting vaccination status and hospitalisation due to influenza-like-illness should have been measured. The second relates to positivity, each individual should have a non-zero probability of getting the high-dose influenza vaccine. Additionally, the propensity score model should be correctly defined, and there should not be any interference between individuals, i.e., an individual vaccinated with the high-dose influenza vaccine will not affect the hospitalisation of another individual. This final assumption has implications in estimating absolute IVE studies due to herd immunity since vaccinated individuals could indirectly protect unvaccinated individuals, leading to IVE underestimation. To account for potential violations of this assumption, we will consider statistical methods that account for clustering, which could be the geographical region of residence.

The propensity score will be estimated using logistic regression, where the vaccination status will be the outcome, and the baseline individual characteristics will be used to adjust. Then, the vaccinated with high-dose influenza vaccine and vaccinated (rIVE) or unvaccinated (absolute IVE) individuals with similar propensity scores will be matched, using nearest neighbour matching, and compared to ensure the balance of covariates. Once the balanced dataset is obtained, the Cox proportional hazards regression model will be used, with the adjustments mentioned in the section above to adjust for residual confounding (13).

### Ethics

This protocol was submitted to the National Institute of Health Doutor Ricardo Jorge Ethics Committee (INSA-EC) in 2024 July 19^th^ and approved on July 30^th^.

## Discussion

This article outlines the conceptual framework and methodological approach to estimate high-dose quadrivalent IVE using EHR in the community and LTCF. We intend to estimate the risk of hospitalisation reduction by comparing two vaccination strategies (High-dose quadrivalent influenza vaccine and standard-dose influenza vaccine – rIVE), and comparing vaccinated with unvaccinated individuals (absolute IVE). To date, retrospective cohort studies regarding high-dose quadrivalent IVE in reducing influenza-related hospitalisation of individuals aged 65 or older have been conducted using specific populations, such as United States veterans or health insurance beneficiaries (10–13). To the best of our knowledge, studies on the effect of high-dose influenza vaccines in LTCF remain limited to immunogenicity studies.

Given the population under study, LTCF and community-dwelling individuals (≥85 years), the potential low vaccine coverage in the eligible population, and the absence of specific studies designed to collect data in LTCF settings, we decided to use routinely collected data in electronic health registries to estimate the high-dose quadrivalent IVE. This approach has already been used successfully to estimate COVID-19 VE (14), and allows VE monitoring with good precision and high representativeness(15–18).

Despite its strengths, this study also has some limitations. We plan to use EHR, which does not aim to collect data for research purposes. Thus, this could lead to misclassification bias regarding vaccine status, outcomes, and confounding variables. For this reason, we might be unable to control for residual confounding since many relevant variables are not monitored or available in registries, and one should be aware that estimates can vary in the presence of confounding. Additionally, unvaccinated individuals will be the reference group when estimating absolute IVE. However, these individuals might differ from the vaccinated individuals due to their clinical profile or possible misclassification bias. Thus, we will use propensity score matching to account for this, assigning a probability of being vaccinated to each individual based on covariates. Considering that we will use EHRs, we might also be unable to correctly identify all individuals living in LTCF, as the report might vary geographically and temporally. Additionally, identifying these individuals might be associated with a differential bias, assuming that vaccinated individuals might be more likely to be reported as living in LTCF than unvaccinated. This situation is less likely to be problematic when estimating relative and absolute IVE in the community.

Another limitation is the potential outcome misclassification. In VE studies, it is extremely important to have sensible outcome definitions as the vaccines are designed to protect against influenza and related complications and studies use laboratory-confirmed outcomes. Using specific influenza ICD codes to identify severe influenza could underestimate severe influenza as the diagnosis is highly dependent on the testing strategy and, thus, on the codification attributed at discharge. On the other hand, including a broader range of ICD codes in the outcome definition could also underestimate IVE by including non-influenza-related outcomes. To balance this, we intend to use ICD codes that have been used for a long time in influenza surveillance and are highly sensitive. Finally, we intend to use discharge codes, as they are the final primary diagnosis of the hospitalisation. However, there are delays in hospital discharge and the codification of the hospitalisation event. In order to minimise this, the retrospective study will only use end-of-season data.

In conclusion, the study aims to produce estimates of high-dose IVE in the population eligible for the high-dose vaccine and contribute to the overall knowledge of the potential added protection provided by this vaccine. Given the vaccination strategy implemented in this population and the national effort in acquiring and implementing vaccination strategies, evaluating the effect of such public health intervention is crucial. The results will also inform decision-makers in future IVE. Additional future studies should include, not only relative indicators, but also quantitative measures of the vaccination strategy’s impact on the population (19).

## Data Availability

No datasets were generated or analysed during the current study. Upon study completion, data cannot be shared publicly because of data confidentiality issues.

## Authors’ contributions

PS and AM conceptualised and designed the study. AM, PS and VG wrote the first draft of the protocol. APR, VG, JAS provided technical inputs. All authors revised and approved the final version.

